# Whole-genome sequence genome-wide association study in *All of Us* identifies a novel glaucoma risk locus in African ancestry individuals

**DOI:** 10.64898/2026.03.19.26348739

**Authors:** Inas F. Aboobakar, Lauren A. Cruz, Tyler G. Kinzy, Yuyang Luo, Sanjana Nallapaneni, Ron Do, Thi Ha Vy, Hetince Zhao, Jessica Tran, Pirro G. Hysi, Anthony P. Khawaja, Puya Gharahkhani, Louis R. Pasquale, Michael A. Hauser, International Glaucoma Genetics Consortium, Ayellet V. Segrè, Dana C. Crawford, Janey L. Wiggs, Jessica N. Cooke Bailey

**Author notes:** These authors contributed equally (Inas F. Aboobakar, Lauren A. Cruz). These authors jointly supervised the work and are co-senior/supervising authors (Janey L. Wiggs, Jessica N. Cooke Bailey). **Address for reprints**: Jessica N. Cooke Bailey, PhD, MA, East Carolina University, 600 Moye Blvd, BSOM 6E 120C, Greenville, NC 27834. **Meeting Presentation:** American Glaucoma Society 2025 Meeting (Top 5 Plenary Paper). **Financial Support:** The study presented in this manuscript was funded by the National Institutes of Health (K23EY035734 to IFA, 5T32EY007157 and 5T32HL007567 to LAC, R01EY022305 to JLW, R01EY033829 to JNCB, R01EY031424 to AVS, R01EY032559 to LRP and JLW, R01EY036460 to LRP, and P30EY014104 to Massachusetts Eye and Ear), Research to Prevent Blindness (Tom Wertheimer Career Development Award in Data Science to IFA), American Glaucoma Society (MAPS Award and Young Clinician-Scientist Grant to IFA), Prevent Blindness Ohio (Young Investigator Student Fellowship Award for Female Scholar in Vision Research to LAC), The Glaucoma Foundation (LRP), UK Research and Innovation Future Leaders Fellowship (MR/Y033930/1 to APK), Alcon Research Institute (Young Investigator Award to APK), Lister Institute for Preventive Medicine Award (APK), NIHR Biomedical Research Centre at Moorfields Eye Hospital (APK), and UCL Institute of Ophthalmology (APK). The sponsor or funding organizations had no role in the design or conduct of this research.

## Abstract

**Objective:** To assess how whole genome sequencing and varying phenotype definitions influence genetic discovery for primary open-angle glaucoma (POAG) in a diverse population.

**Design:** Ancestry-stratified genome-wide association studies (GWASs) and cross-ancestry meta-analyses of POAG cases and controls using two phenotype definitions.

**Participants:** Cases (age≥40) and controls (age≥65) were identified in the National Institutes of Health *All of Us* Research Program v8 data release and sub-divided into genetically inferred ancestral groups. Using the relaxed phenotype (ICD codes only), case/control counts were: European (1,846/84,654), African (1,042/15,966), and Latino/Admixed American (305/10,167). Using the stringent phenotype (ICD codes and evidence of glaucoma treatment in the electronic health record), case/control counts were: European (1,528/79,276), African (862/14,076), and Latino/Admixed American (250/9,668). Cross-ancestry meta-analyses included 3,193 cases/110,787 controls for the relaxed phenotype and 2,640 cases/103,020 controls for the stringent phenotype.

**Methods:** GWASs were conducted within European, African, and Latino/Admixed American ancestry groups individually using firth logistic regression with age, sex, and the top 10 genotype principal components included as covariates. The ancestry-stratified GWASs were then meta-analyzed using a fixed-effects, inverse variance-weighted approach.

**Main Outcome Measures:** Identification of genome-wide significant loci (*P <* 5×10^-8^) for POAG using different phenotype definitions and ancestry groups.

**Results:** Known POAG risk loci (e.g., *TMCO1*, *CDKN2B-AS1*, and *GMDS*) reached genome-wide significance in both the European GWASs and cross-ancestry meta-analyses (odds ratio (OR) range: 1.19–1.38). A novel risk locus near *CYP2A7* (rs76935404[T], OR = 1.35) was identified in the African ancestry GWAS using the stringent phenotype definition. Effect sizes for known POAG risk loci from prior large-scale meta-analyses strongly correlated with effect sizes in this study (Pearson r = 0.75–0.84, *P <* 1 × 10⁻⁵ for all). The strength and consistency of these correlations support the robustness of the findings.

**Conclusions:** This study demonstrates the value of whole genome sequencing, diverse ancestry inclusion, and phenotypic refinement in uncovering novel POAG genetic risk loci. The findings underscore the need to prioritize both genetic diversity and refined case/control definitions to advance understanding of this complex ocular disease.

**Précis:** This study identifies a novel primary open-angle glaucoma risk locus in individuals of African ancestry using whole genome sequencing and varying phenotype definitions in the diverse *All of Us* Research Program dataset.

---

While primary open-angle glaucoma (POAG) is prevalent in all populations, population-based studies suggest it disproportionately affects African and Latino individuals, who have a younger age at onset, present at more advanced disease stages, and have greater risk of blindness compared to non-Hispanic whites.^1–5^ More than 300 risk loci have been identified in prior analyses of this highly heritable complex disease^6–8^, which have mainly been conducted in individuals of European ancestry (EUR). However, few are conclusively associated with risk of glaucoma in individuals of African continental genetic ancestry (AFR)^9–14^, and even fewer in those with Latino/Admixed ancestry (AMR).^10,14^

In addition to the limited ancestral diversity in prior POAG studies, most known loci have been identified in genome-wide association studies (GWASs) conducted on genotyping arrays, where only a subset of variants (∼2%) across the genome are directly assayed.^15^ The remaining variants examined in these GWASs are typically imputed, requiring large reference panels of sequenced individuals of similar ancestry to the population being studied.^16^ Imputation accuracy is highly variable and especially dependent on the ancestry of the reference and genotyped population samples.^17^ In contrast, whole genome sequencing (WGS) can provide more comprehensive ascertainment of both common and rare putative causal variants in genetic risk loci.^18^

Phenotype definitions also vary widely across genetic studies, ranging from self-reported diagnosis or ICD-9/10 codes alone to comprehensive clinical examinations by ophthalmologists. Studies using large-scale biobanks as a sample source often rely on ICD diagnosis codes for disease ascertainment, as ocular exam and imaging data are often limited.^19^ However, ICD codes as phenotype definitions may have reduced specificity and potentially greater heterogeneity, diminishing statistical power to detect genetic differences between individuals with and without a disease of interest.

The National Institutes of Health *All of Us* (*AoU*) Research Program dataset enables examination of the impact of phenotype variability and WGS on genomic discovery for complex-inherited diseases such as POAG. *AoU* aims to enroll over one million individuals of diverse backgrounds to improve disease diagnosis, treatment, and prevention.^20^ Data from surveys, WGS, electronic health records (EHRs), physical measurements, and wearable devices are available in the *AoU* Researcher Workbench.^20,21^ To date, WGS data have been released for over 414,000 participants (v8), nearly half of whom are from groups historically underrepresented in biomedical research.^21^ In this study, we apply two phenotype definitions (relaxed and stringent) for POAG cases and controls in *AoU*. We then perform ancestry-stratified GWASs and cross-ancestry meta-analyses of EUR, AFR, AMR individuals using WGS data. Using this approach, we have replicated known POAG risk loci and identified a novel risk locus in AFR individuals.

## Methods

An overview of the study design is shown in **Figure 1**. Informed consent was obtained for all participants enrolled in *AoU*. The *AoU* IRB follows the regulations and guidance of the National Institutes of Health Office for Human Research Protections for all studies, ensuring that the rights and welfare of research participants are overseen and protected uniformly.^20,21^ The described research adhered to the tenets of the Declaration of Helsinki.

**Figure 1.**
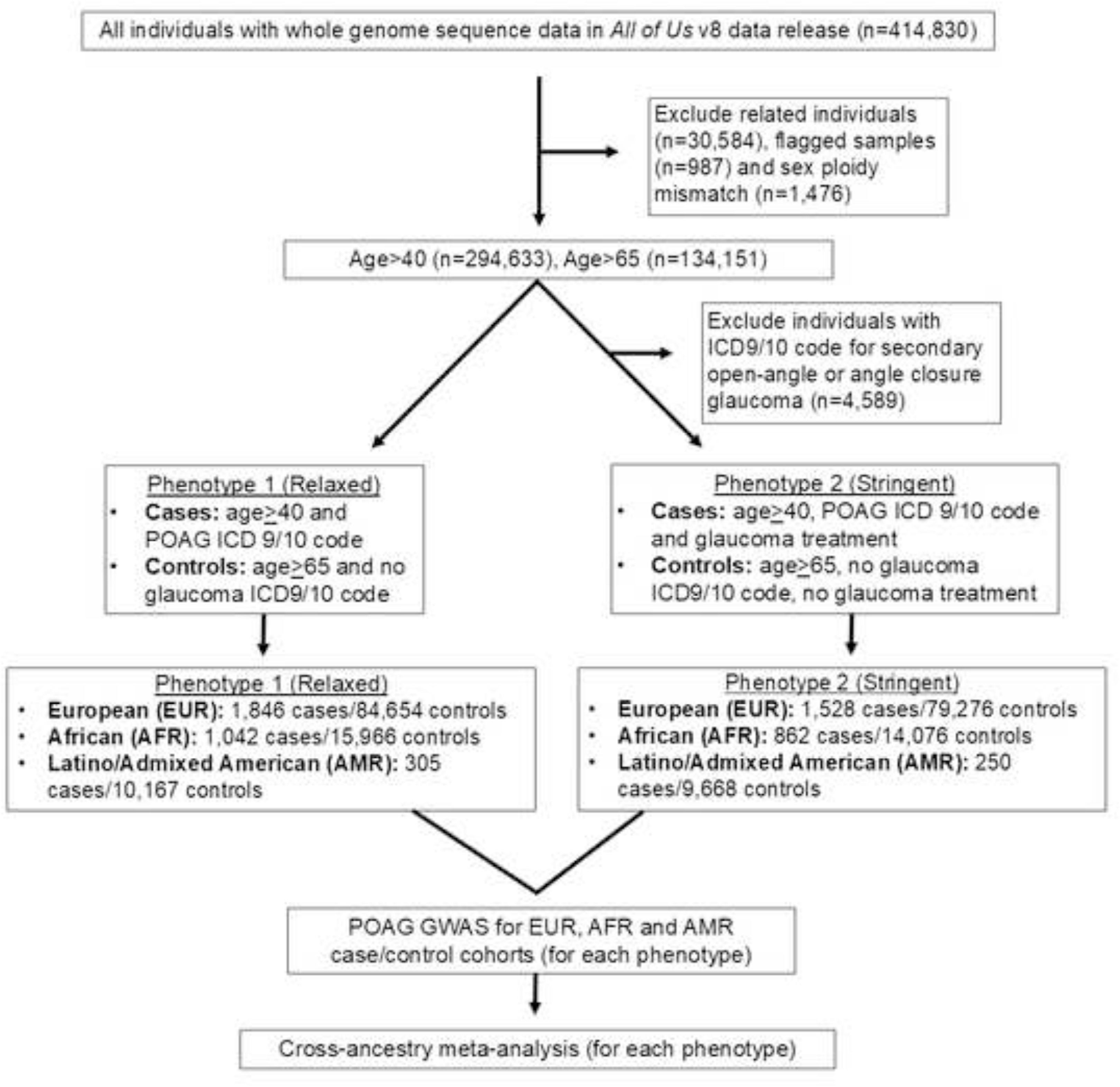
Study design. Abbreviations used: POAG (primary open-angle glaucoma); GWAS (genome-wide association study).

### Case/control definitions

Our baseline inclusion criteria were as follows: *AoU* participants with WGS data and current age ≥40 at the time of analysis (February 2025), which were identified using the cloud-based *AoU* Researcher Workbench. For Phenotype 1 (relaxed), POAG cases were defined as individuals age≥40 with an ICD-9/10 diagnosis code for POAG, but no code for other secondary forms of open-angle glaucoma or primary/secondary angle closure glaucoma (**Supplementary Table 1**). Controls were age≥65 and did not have any ICD-9/10 diagnosis codes for POAG, any other form of glaucoma, glaucoma suspect, ocular hypertension, or family history of glaucoma; an older cohort was used for controls to minimize the likelihood of undiagnosed cases in the control dataset. For Phenotype 2 (stringent), POAG cases met criteria for Phenotype 1 and also had evidence of glaucoma treatment (topical intraocular pressure lowering drops, laser trabeculoplasty, minimally invasive glaucoma surgery, or incisional glaucoma surgery) via RxNorm and CPT codes in the EHR; controls met the criteria for Phenotype 1 and also did not have any evidence of glaucoma treatment in their EHR (**Supplementary Table 1**).

### Whole genome sequencing data and quality control

The *AoU* WGS protocol has been extensively described previously.^21^ Briefly, WGS was performed on the Illumina NovaSeq 6000 instrument, and analyses were performed on the Illumina DRAGEN platform. Quality control (QC) metrics included the following: mean coverage (threshold of ≥30×), genome coverage (threshold of ≥90% at 20×), coverage of hereditary disease risk genes (threshold of ≥95% at 20×), aligned Q30 bases (threshold of ≥8 × 10^10^), contamination (threshold of ≤1%), and concordance to independently processed array data.^21^ Additional sample QC metrics were also applied by *AoU*, including fingerprint concordance, sex concordance, and cross-individual contamination rate. WGS from 414,830 participants passed all QC metrics and are available in the *AoU* v8 data release. All variants reported as genome-wide significant passed standard *All of Us* v8 variant-level quality control filters, including thresholds for call rate, Hardy–Weinberg equilibrium (HWE), and minor allele frequency (MAF). Specifically, variant quality control was performed starting with 116,456,419 SNPs, and missingness<0.02. After all QC steps, under the Phenotype 1 (relaxed) POAG definition, 9,890,817 variants remained in the EUR group, 17,099,446 in the AFR, and 11,153,168 in the AMR. Under the Phenotype 2 (stringent) POAG definition, 9,893,120 variants (8.5%) remained in the EUR ancestry group, 17,097,654 (14.7%) in the AFR, and 11,308,763 (9.7%) in the AMR.

Genetically-inferred ancestry was derived by *AoU* for all participants with WGS data in the v8 data release; the ancestry labels are consistent with those in Genome Aggregation Database (gnomAD), the Human Genome Diversity Project, and 1000 Genomes.^21^ Individuals with kinship scores >0.1 suggesting relatedness (n=30,584), ancestral outliers on principal components analysis (n=987), or sex ploidy mismatch (n=1,476) were excluded from the POAG GWASs.

### Ancestry-stratified GWASs and cross-ancestry meta-analyses

Firth logistic regression analyses were performed using HAIL version 0.2.134 for each ancestral group individually for each phenotype definition, with age, sex, and the top 10 genotype principal components included as covariates.

Genetic heterogeneity across the individual GWASs were assessed using the I^2^ test in METAL; there was no evidence of significant heterogeneity in the EUR, AFR, and AMR GWASs.^22^ The summary statistics for each phenotype definition were meta-analyzed using a fixed-effects, inverse variance-weighted approach in METAL (release date 2011-03-25) (3,193 cases and 110,787 controls for Phenotype 1; 2,640 cases and 103,020 controls for Phenotype 2).^22^ To ensure that a random-effects model would yield a consistent result, the GWAS summary statistics were also meta-analyzed using MR-MEGA.^23^ Manhattan plots, quantile-quantile (QQ) plots, and LocusZoom plots were created in LocusZoom.js (v0.12).^24^

### Correlation of effect sizes for known POAG variants

The effect sizes (beta coefficients) of POAG-associated variants identified in previously reported cross-ancestry meta-analyses, including the International Glaucoma Genetics Consortium (IGGC) meta-analysis (127 variants)^7^ and the multi-trait analysis of GWAS (MTAG) meta-analysis (312 variants)^8^ were compared to the *AoU* POAG cross-ancestry meta-analyses for both the relaxed and stringent phenotype definitions. Additionally, the effect sizes for POAG variants identified in the IGGC EUR GWAS meta-analysis (68 variants)^7^ were compared to their effect sizes in the *AoU* EUR GWASs. The correlation between effect sizes was assessed using Pearson’s correlation coefficient (r).

### Cross-reference of the novel genome-wide significant locus in an external dataset

To evaluate the robustness of the novel genome-wide significant variant (rs76935404[T] near *CYP2A7*), we queried this variant in an independent dataset from the Genetics of Glaucoma in People of African Descent (GGLAD) Consortium recruited at Duke University (Durham, NC, USA). The GGLAD-Duke dataset consists of genotype array data from 701 cases and 606 controls, as previously described.^11^ The association statistics for rs76935404 were derived using the same phenotype definition of primary open-angle glaucoma and standard GWAS quality control procedures applied in the original study. After nominal replication was noted, the GGLAD-Duke summary statistics were meta-analyzed with the AFR GWAS summary statistics for each phenotype definition using a fixed-effects, inverse variance-weighted approach in METAL.^22^

### *In-silico* annotation

To assess whether any novel genome-wide significant variants were expression quantitative trait loci (eQTLs) that could regulate the expression of neighboring or distant genes, these variants were queried in the Gene-Tissue Expression (GTEx) database^25^ and the University of Michigan’s FIVEx^26^ browser. Because ocular tissue expression data are not currently available in existing databases, we used other neuronal tissues as a surrogate, given that POAG is also a neurodegenerative disease. The Broad Institute Single Cell Portal^27^ was queried to identify ocular cell types in which genes located within novel risk loci are expressed.

## Results

### POAG GWASs in the *AoU* European (EUR), African (AFR), and Latino/Admixed American (AMR) cohorts

Demographic characteristics of the ancestry-specific cohorts are summarized in **Supplementary Table 2**. Results from the EUR (1,846 vs. 1,528 cases/84,654 vs. 79,276 controls), AFR (1,042 vs. 862 cases/15,966 vs. 14,076 controls), and AMR (305 vs. 250 cases/10,167 vs. 9,668 controls) GWASs for each phenotype definition are shown in **Figure 2** and **Table 1**. There was no evidence of significant genomic inflation or deflation in any of these GWASs (lambda=1.0-1.02) (**Supplementary Figure 1 A-C**). Under both phenotype definitions, in AFR and AMR subsets, but not in EUR, cases were significantly younger than controls. Significant sex differences were observed, with controls more likely to be female than male in all groups, except for the AFR group (under both phenotype definitions) and the AMR group (from Phenotype 1), where the proportion of females did not differ significantly between cases and controls. For Phenotype 1 (relaxed), two loci reached genome-wide significance (*P <* 5×10^-8^) in the EUR GWAS, both of which are known POAG risk loci: *TMCO1* (rs6426939[C], OR = 1.32, *P =* 1.7×10^-8^) and *CDKN2B-AS1* (rs3217977[C], OR = 1.21, *P =* 3.8×10^-8^). No significant loci were detected in the AFR or AMR GWASs. For Phenotype 2 (stringent), one locus reached genome-wide significance in the EUR GWAS, which is also a known locus: *GMDS* (rs1570537[C], OR = 1.38, *P =* 1.0×10^-8^). No significant loci were identified in the AMR GWAS. Interestingly, a novel risk variant near *CYP2A7* was identified in the AFR GWAS (rs76935404[T], OR = 1.35, *P =* 1.2×10^-8^). LocusZoom plots for genome-wide significant loci are shown in **Supplementary Figure 2**.

**Figure 2.**
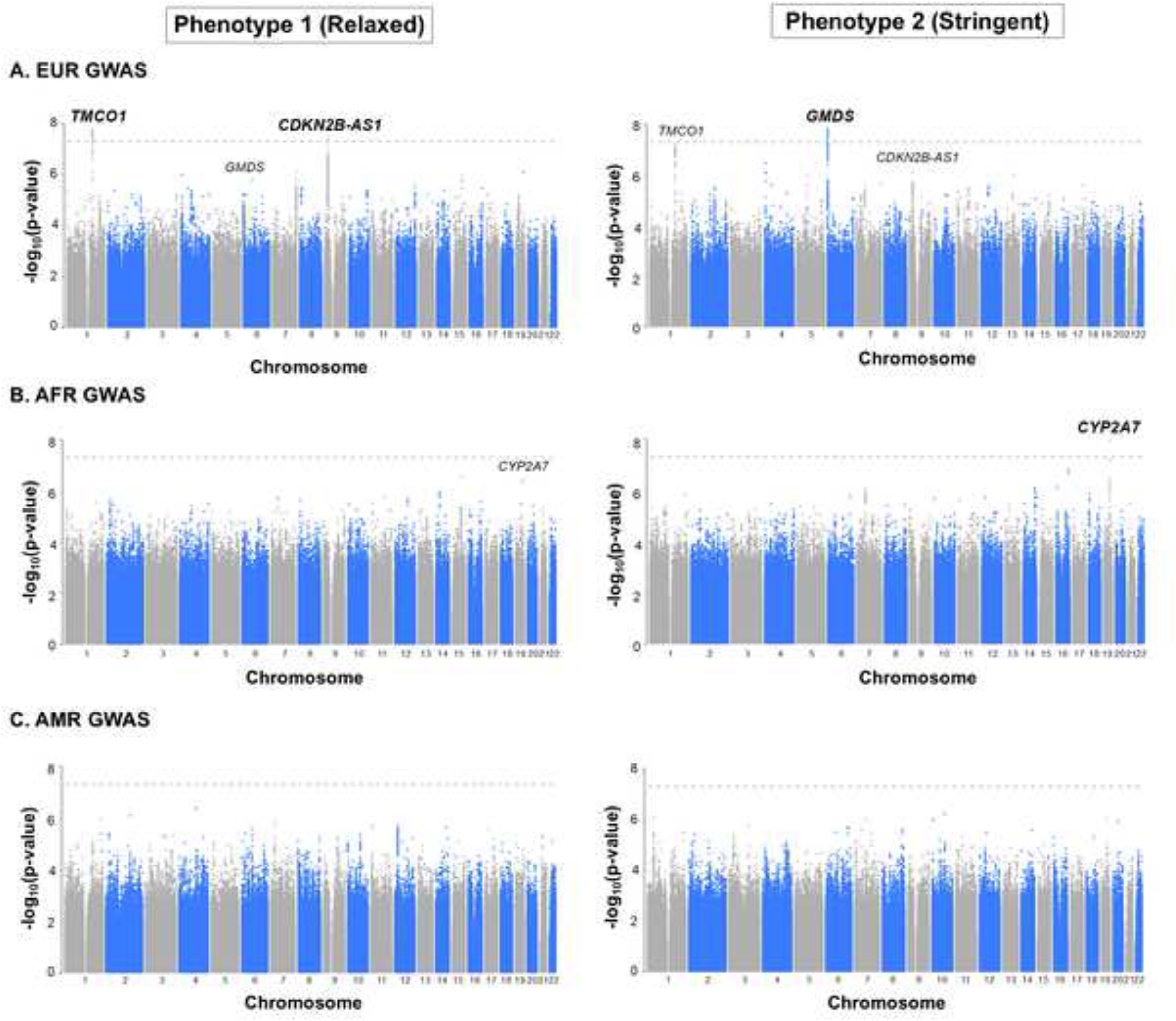
Manhattan plots for primary open-angle glaucoma (POAG) ancestry-stratified genome-wide association studies (GWASs) in the *All of Us* Research Program (A) European (EUR), (B) African (AFR), and (C) Latino/Admixed American (AMR) cohorts using each phenotype definition (relaxed and stringent). Each dot represents a specific variant. The x-axis displays the chromosome where each variant is located, and the y-axis displays the -log_10_ *P*-value for association of the variant with POAG. The grey dashed line represents the genome-wide significance threshold (*P =* 5×10^-8^, -log_10_p = 7.30). For variants reaching genome-wide significance, the nearest gene to the lead variant in the locus is labeled and bolded. The nearest gene(s) to genome-wide significant loci using one phenotype definition are also labeled on the respective Manhattan plots for the second phenotype definition.

**Table 1.** Genome-wide significant variants in the *All of Us* ancestry-specific POAG GWASs using each phenotype definition.

| GWAS | Cases/Controls | Chr | Position (GRCh38) | Lead Variant | Nearest Gene | Effect Allele | Other Allele | OR | 95% CI | P-value | Case EAF | Control EAF | Known or novel locus |
| --- | --- | --- | --- | --- | --- | --- | --- | --- | --- | --- | --- | --- | --- |
| EUR | 1,846/84,654 | 1 | 165746063 | rs6426939 | *TMCO1* | C | T | 1.32 | 1.20-1.45 | 1.7x10<sup>-8</sup> | 0.15 | 0.12 | Known |
|  |  | 9 | 22007358 | rs3217977 | *CDKN2B-AS1* | C | CA | 1.21 | 1.13-1.29 | 3.8x10<sup>-8</sup> | 0.60 | 0.55 | Known |

Phenotype 2 (Stringent)
| GWAS | Cases/Controls | Chr | Position (GRCh38) | Lead Variant | Nearest Gene | Effect Allele | Other Allele | OR | 95% CI | P-value | Case EAF | Control EAF | Known or novel locus |
| --- | --- | --- | --- | --- | --- | --- | --- | --- | --- | --- | --- | --- | --- |
| EUR | 1,528/79,276 | 6 | 1962530 | rs1570537 | *GMDS* | C | T | 1.38 | 1.24-1.55 | 1.0x10<sup>-8</sup> | 0.89 | 0.86 | Known |
| AFR | 862/14,076 | 19 | 40913389 | rs76935404 | *CYP2A7* | T | C | 1.35 | 1.22-1.50 | 1.2x10<sup>-8</sup> | 0.35 | 0.28 | Novel |

POAG: primary open-angle glaucoma; GWAS: genome-wide association study; EUR: European; AFR: African; Chr: chromosome; OR: odds ratio; CI: confidence interval; EAF: effect allele frequency (in specific population in which GWAS was performed)

The novel variant rs76935404 is found in a non-coding region of the genome upstream of the promoter for the *CYP2A7* gene. As non-coding variants can regulate the expression of both neighboring and distant genes^28^, we queried whether this variant is an expression quantitative trait locus (eQTL) and thereby influences gene expression. Because existing databases do not contain ocular tissue expression data, neuronal tissues were used as a surrogate given POAG is also a neurodegenerative disease. rs76935404 was found to be a significant eQTL for: 1) *CYP2A7* in the brain hypothalamus, basal ganglia, and hippocampus; 2) *EGLN2* in sensory neuronal tissue; and 3) the uncharacterized long non-coding RNA AC092071.1 in the dorsal prefrontal cortex (*P <* 0.05 for all). Intriguingly, the coding genes have POAG-relevant biological functions: *CYP2A7*^29,30^ is a member of the Cytochrome P450 superfamily of enzymes that function in drug metabolism and steroid/lipid biosynthesis, while *EGLN2*^31–33^ plays roles in oxygen sensing, apoptosis and mitochondrial homeostasis. Single-cell data also shows detectable expression of both genes in disease-relevant ocular tissues from healthy donor eyes.^27,34^ *CYP2A7* is expressed in optic nerve astrocytes, lens, Müller glia, and retinal ganglion cells, while *EGLN2* is expressed in optic nerve astrocytes, lens, and optic nerve head fibroblasts.

To further validate the novel variant rs76935404 identified in the Phenotype 2 (stringent) AFR GWAS, we queried this variant in a subset of the Genetics of Glaucoma in People of African Descent (GGLAD) Consortium recruited at Duke University (Durham, NC, USA) (GGLAD-Duke) after a comprehensive ocular examination by an ophthalmologist (701 cases/606 controls).^11^ rs76935404 replicated with same direction of effect and nominal significance in the GGLAD-Duke dataset (OR = 1.34, *P =* 5.8×10^-3^) (**Table 2**). Given there was evidence of replication, we meta-analyzed the GGLAD-Duke summary statistics with the *AoU* AFR GWASs for each phenotype definition using a fixed-effects, inverse-variance weighted approach. Meta-analyses showed that rs76935404 reached genome-wide significance using the relaxed phenotype definition (OR = 1.29, *P =* 9.6×10^-9^) and maintained significance using the stringent definition (OR = 1.35, *P =* 2.3×10^-10^) (**Supplementary Figure 3**). These meta-analyses did not show any evidence of significant genomic inflation (λ = 1.02-1.03) (**Supplementary Figure 4**).

**Table 2.**
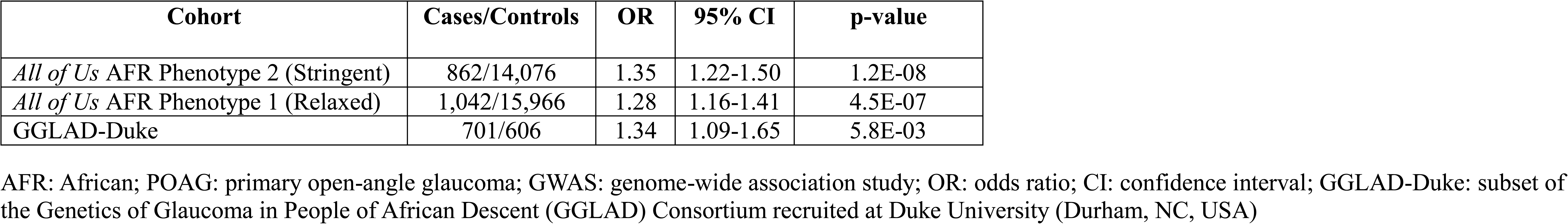
Association of the novel genome-wide significant variant from the *All of Us* AFR Phenotype 2 (Stringent) POAG GWAS (rs76935404[T]) in an independent cohort.

### Cross-ancestry POAG GWAS meta-analyses of the *All of Us* EUR, AFR, and AMR cohorts

The known POAG risk loci *TMCO1* (rs35862498[A], OR = 1.23, *P =* 1.1×10^-9^) and *CDKN2B-AS1* (rs3217977[C], OR = 1.19, *P =* 5.9×10^-9^) were genome-wide significant in a fixed effects, inverse variance-weighted cross-ancestry meta-analysis using Phenotype 1 (relaxed) (3,193 cases /110,787 controls), while only *TMCO1* (Chr1:165730355[CA], OR = 1.26, *P =* 3.5×10^-9^) was significant using Phenotype 2 (stringent) with a reduced cohort size of 2,640 cases and 103,020 controls (**Figure 3**, **Table 3**, **Supplementary Figure 5**). Neither GWAS meta-analysis demonstrated evidence of genomic inflation (λ = 1.0-1.01) (**Supplementary Figure 1D**). To ensure the results would be consistent with a random effects model, the meta-analyses were repeated with this approach, which yielded the same genome-wide significant loci (**Supplementary Figure 6**).

**Figure 3.**
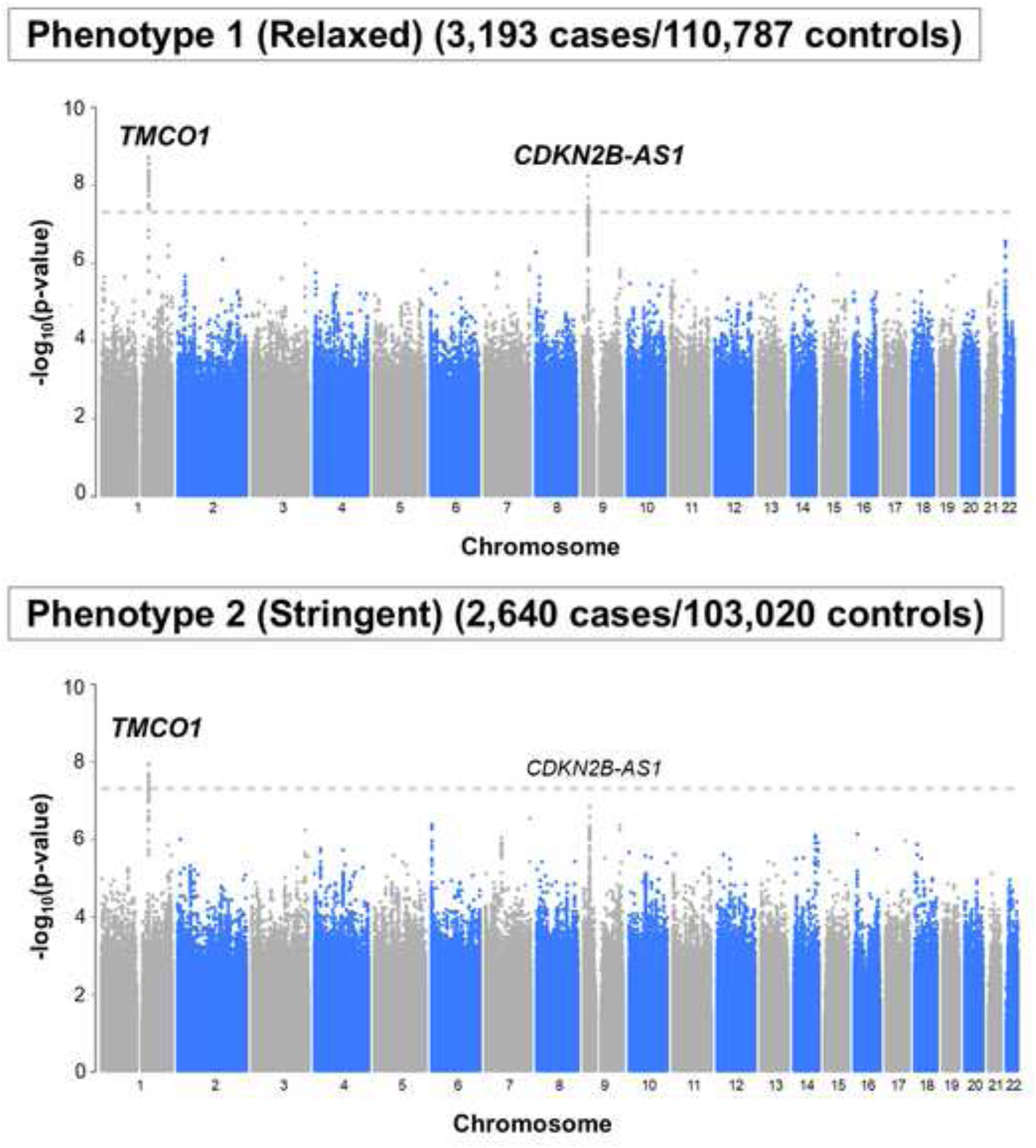
Manhattan plots for the *All of Us* Research Program cross-ancestry primary open-angle glaucoma (POAG) genome-wide association study (GWAS) meta-analyses using each phenotype definition (relaxed and stringent). GWASs were performed for the European (EUR), African (AFR), and Latino/Admixed American (AMR) cohorts using a relaxed and stringent phenotype (Figure 2) and the results were then meta-analyzed for each phenotype definition. Each dot represents a specific variant. The x-axis displays the chromosome where each variant is located, and the y-axis displays the -log_10_ *P*-value for association of the variant with POAG in the respective cross-ancestry meta-analysis. The grey dashed line represents the genome-wide significance threshold (*P =* 5×10^-8^, -log_10_p = 7.30). For variants reaching genome-wide significance, the nearest gene to the lead variant in the locus is labeled and bolded. The nearest gene(s) to genome-wide significant loci in one phenotype definition are also labeled on the Manhattan plot for the second phenotype definition.

**Table 3.**
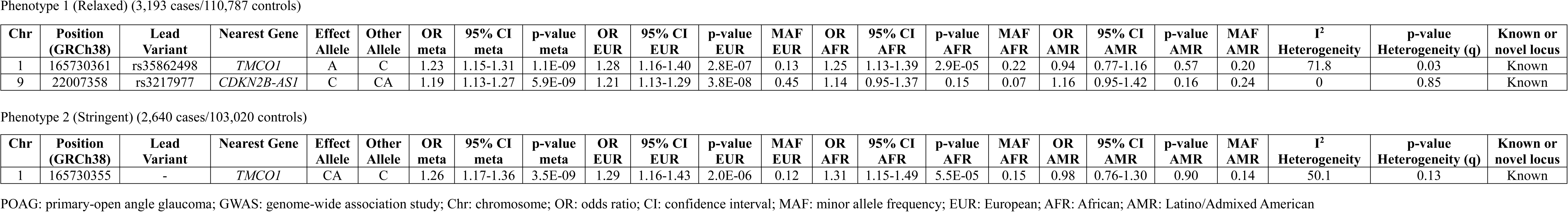
Genome-wide significant variants in the *All of Us* cross-ancestry POAG GWAS meta-analyses using each phenotype definition.

Phenotype 1 (Relaxed) (3,193 cases/110,787 controls)
| Chr | Position (GRCh38) | Lead Variant | Nearest Gene | Effect Allele | Other Allele | OR meta | 95% CI meta | p-value meta | OR EUR | 95% CI EUR | p-value EUR | MAF EUR | OR AFR | 95% CI AFR | p-value AFR | MAF AFR | OR AMR | 95% CI AMR | p-value AMR | MAF AMR | I<sup>2</sup> Heterogeneity | p-value Heterogeneity (q) | Known or novel locus |
| --- | --- | --- | --- | --- | --- | --- | --- | --- | --- | --- | --- | --- | --- | --- | --- | --- | --- | --- | --- | --- | --- | --- | --- |
| 1 | 165730361 | rs35862498 | *TMCO1* | A | C | 1.23 | 1.15-1.31 | 1.1E-09 | 1.28 | 1.16-1.40 | 2.8E-07 | 0.13 | 1.25 | 1.13-1.39 | 2.9E-05 | 0.22 | 0.94 | 0.77-1.16 | 0.57 | 0.20 | 71.8 | 0.03 | Known |
| 9 | 22007358 | rs3217977 | *CDKN2B-AS1* | C | CA | 1.19 | 1.13-1.27 | 5.9E-09 | 1.21 | 1.13-1.29 | 3.8E-08 | 0.45 | 1.14 | 0.95-1.37 | 0.15 | 0.07 | 1.16 | 0.95-1.42 | 0.16 | 0.24 | 0 | 0.85 | Known |

Phenotype 2 (Stringent) (2,640 cases/103,020 controls)
| Chr | Position (GRCh38) | Lead Variant | Nearest Gene | Effect Allele | Other Allele | OR meta | 95% CI meta | p-value meta | OR EUR | 95% CI EUR | p-value EUR | MAF EUR | OR AFR | 95% CI AFR | p-value AFR | MAF AFR | OR AMR | 95% CI AMR | p-value AMR | MAF AMR | I<sup>2</sup> Heterogeneity | p-value Heterogeneity (q) | Known or novel locus |
| --- | --- | --- | --- | --- | --- | --- | --- | --- | --- | --- | --- | --- | --- | --- | --- | --- | --- | --- | --- | --- | --- | --- | --- |
| 1 | 165730355 | - | *TMCO1* | CA | C | 1.26 | 1.17-1.36 | 3.5E-09 | 1.29 | 1.16-1.43 | 2.0E-06 | 0.12 | 1.31 | 1.15-1.49 | 5.5E-05 | 0.15 | 0.98 | 0.76-1.30 | 0.90 | 0.14 | 50.1 | 0.13 | Known |

POAG: primary-open angle glaucoma; GWAS: genome-wide association study; Chr: chromosome; OR: odds ratio; CI: confidence interval; MAF: minor allele frequency; EUR: European; AFR: African; AMR: Latino/Admixed American

The novel risk locus identified in the AFR Phenotype 2 GWAS (rs76935404 near *CYP2A7*) was polymorphic in and exhibited a similar direction of effect across all ancestries, though the strength of effect was strongest in the AFR cohort; consequently, this locus did not remain significant in the cross-ancestry meta-analyses (OR = 1.09-1.11, *P <* 5×10^-3^ for all) (**Supplementary Table 3**).

### Correlation of published effect sizes at known POAG risk loci with effect sizes in the *AoU* meta-analysis

As measures of data quality, validity, and to evaluate the utility for additional genetic studies of POAG, we compared the effect sizes (beta regression coefficients) for known POAG risk loci with their effect sizes in the *AoU* GWASs. **Figure 4A** shows the effect size comparisons of 127 variants identified in the International Glaucoma Genetics Consortium (IGGC POAG cross-ancestry meta-analysis (34,179 cases/349,321 controls).^7^ **Figure 4B** shows the effect size comparisons of 312 variants identified in the multi-trait analysis of GWAS (MTAG) POAG cross-ancestry meta-analysis (39,457 cases/392,560 controls).^8^ For the 127 IGGC variants as well as the 312 MTAG variants, there was strong correlation between effect sizes from the prior analyses and the *AoU* meta-analyses using both phenotype definitions (r=0.83-0.84 for IGGC variants, *P <* 1×10^-5^; r=0.75-0.79 for MTAG variants, *P <* 1×10^-5^, respectively). For the 68 variants previously identified in the IGGC EUR POAG meta-analysis (16,677 cases/199,580 controls)^7^, the correlation between IGGC EUR effect sizes and the *AoU* EUR effect sizes using both phenotype definitions was also strong (r=0.86-0.89, *P <* 1×10^-5^; **Supplementary Figure 7**). For internal validation, the effect sizes of top variants (*P <* 1×10^-5^) from ancestry-stratified and cross-ancestry meta-analyses applying Phenotype 1 and Phenotype 2 were also compared; correlation was again strong (r=0.96-0.99, *P <* 1×10^-5^, **Supplementary Figure 8**).

**Figure 4.**
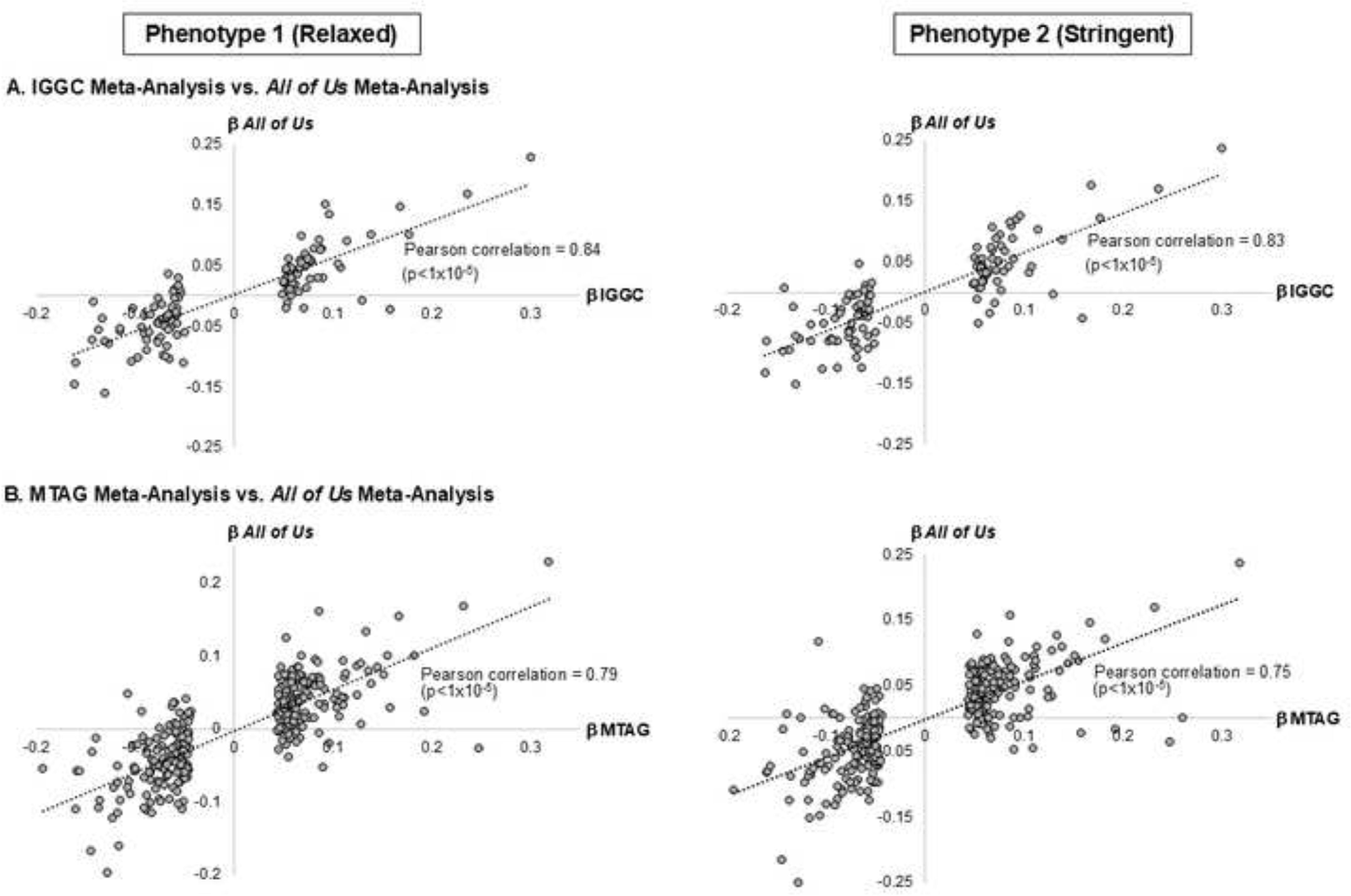
Correlation of effect sizes for known primary open-angle glaucoma (POAG)-associated variants with their effect sizes in the *All of Us* Research Program cross-ancestry meta-analyses using each phenotype definition (relaxed and stringent). Correlation analyses were performed for all POAG-associated variants identified in the (A) International Glaucoma Genetics Consortium (IGGC) POAG cross-ancestry meta-analysis (34,179 cases/349,321 controls; 127 variants)^7^ and the (B) Multi-trait analysis of GWAS (MTAG) POAG cross-ancestry meta-analysis (39,457 cases/392,560 controls; 312 variants).^8^ Each dot represents a specific variant. The beta regression coefficients (ß) from the IGGC/MTAG cross-ancestry meta-analyses are plotted on the x-axis, while the ß from the *All of Us* Research Program cross-ancestry meta-analyses are plotted on the y-axis.

## Discussion

While POAG disproportionately impacts individuals of AFR and AMR ancestry, studies aiming to identify genetic risk factors have predominantly enrolled individuals of EUR ancestry^6^, consistent with the approach for other common diseases.^35^ Additionally, these studies have utilized genotype arrays, which directly assay ∼2% of variants throughout the genome and have historically not been robust to differences in continental genetic ancestry.^36^ Successfully imputing missing genotypes is highly dependent on the size of reference panels, linkage disequilibrium (LD) variation between populations, variant MAF, array marker density, and sequence quality, among other factors.^17^ For samples from populations with more AFR and/or AMR continental genetic ancestry, for which reference panels are smaller than EUR reference panels, accuracy varies widely.^17^ To overcome these shortcomings, this work is the first cross-ancestry GWAS meta-analysis for POAG using WGS data in the ancestrally diverse *AoU* Research Program. In addition, we aimed to overcome the limitations of using ICD codes alone for disease ascertainment by applying different levels of phenotype definition stringency. The combination of these strengths replicated known POAG risk loci and also allowed us to identify a novel AFR risk locus. Moreover, the strong correlation of effect sizes for previously identified variants in the IGGC (r=0.83-84) and MTAG (r=0.75-79) POAG cross-ancestry meta-analyses provides support for data quality and utility of this dataset for future POAG genetic studies.

Prior genome-wide association studies in African ancestry populations have identified relatively few POAG risk loci compared to studies conducted in European ancestry populations. For example, variants near *APBB2* were previously reported as genome-wide significant in African ancestry individuals in the GGLAD Consortium.^11^ In the current study, while previously reported POAG loci were well-represented among sub–genome-wide significant signals, no variants at the *APBB2* locus reached genome-wide significance in the *All of Us* AFR GWAS. Differences in sample size, phenotype definition, study design, and underlying genetic architecture may contribute to variability in locus discovery across studies. These findings underscore the complementary nature of diverse datasets and reinforce the importance of continued large-scale genetic studies in ancestrally diverse populations.

The significant signal observed for rs76935404[T] in the stringently-phenotyped AFR GWAS replicated in the GGLAD-Duke dataset, and the effect size was also consistent with the observation in *AoU* (OR = 1.34). Notably, the variant is polymorphic across ancestry groups but showed the strongest association in individuals of African ancestry, highlighting the importance of including diverse populations in genetic studies of POAG. Stronger effect sizes were detected in datasets with more carefully phenotyped POAG cases. Phenotype 2 in our study considers not only POAG-related ICD 9/10 codes, but also CPT and RxNorm codes for glaucoma treatment to more precisely define POAG cases compared to Phenotype 1. GGLAD-Duke POAG cases and controls were clinically examined for diagnosis.^11^

The novel locus is ∼30 kilobases upstream of the *CYP2A7* gene promoter. *CYP2A7* encodes an orphan cytochrome P450 protein that localizes to the endoplasmic reticulum.^37^ While *CYP2A7* has not been reported in the GWAS Catalog as associated with any ocular phenotypes^38^, another cytochrome P450 gene, *CYP1B1*, has been implicated in primary congenital glaucoma.^39^ Both *CYP2A7* and *CYP1B1* belong to a large cluster of cytochrome P450 genes on chromosome 19q. More targeted high-resolution sequencing interrogation across this region would be needed to confirm a causal variant(s)/gene(s).

While the novel index variant is canonically closest to *CYP2A7*, evidence from the GTEx and FIVEx databases suggest that rs76935404 may also be an eQTL for *EGLN2*, which encodes an enzyme with a crucial role in oxygen sensing. This enzyme catalyzes hydroxylation of specific proline residues in hypoxia-inducible factor alpha (HIF-1ɑ) subunits when oxygen is abundant.^32^ This hydroxylation marks HIF-1ɑ for degradation, effectively regulating cell response to changing oxygen levels. Hypoxia-induced retinal ganglion cell death is implicated in glaucoma pathophysiology.^40^ Interestingly, a recent analysis of rare variants identified using whole exome sequence data reported significant association between variants in *EGLN2* and intraocular pressure.^41^ This study also reported that *EGLN2* is a drug target for three HIF prolyl hydroxylase inhibitors (roxadustat, daprodustat, and vadadustat).

The hypoxia response pathway has been implicated in various adaptive responses to environmental pressures^42^, and variants in oxygen-sensing genes have shown population-specific distributions that may reflect past selective pressures. For example, HIF-1ɑ is induced in host tissues during malaria infection, a disease that has influenced selection pressure of other disease-causing genes (e.x. sickle cell in AFR populations).^43^ Activation of the host HIF-1ɑ pathway improves survival of liver-stage malaria parasites in hepatocytes.^44^ In context, continental AFR populations faced unique selective pressures from endemic diseases such as malaria, sleeping sickness, and various parasitic infections, potentially favoring variants that enhance detoxification capacity or immune responses. The evolutionary maintenance of these variants in AFR populations suggests they may represent ancestry-specific risk alleles that contribute to health disparities in complex diseases. This pattern has been observed for other conditions, such as *APOL1* variants that protect against trypanosomiasis but increase kidney disease risk in African Americans.^45^ Interestingly, in our ancestry-stratified GWASs, the direction of effect for rs76935404[T] was consistent across ancestries, though the effect size was much larger in the AFR GWAS, supporting this theory. Importantly, *CYP2A7* and *EGLN2* expression is detected in disease-relevant ocular tissues from healthy donor eyes (*CYP2A7* is expressed in optic nerve astrocytes, lens, Müller glia, and retinal ganglion cells, while *EGLN2* is expressed in optic nerve astrocytes, lens, and optic nerve head fibroblasts), supporting a potential functional role in POAG pathogenesis. Further functional work is needed to prioritize the causal gene(s) in this locus and is an important future research direction.

A major strength of the current study is the ancestrally diverse population of research participants. This increases statistical power to detect novel POAG susceptibility loci that were previously undiscovered due to lack of representation of non-EUR individuals in most GWASs to date. Another strength of this study is the use of WGS data; imputed array data is currently more commonly used in GWASs than WGS data, though as WGS becomes cheaper and more widely available, more GWASs will likely shift towards WGS data to eliminate the need for imputation and better facilitate harmonization of genomic data collected across different studies/sequencing platforms. Greater use of WGS will also enable discovery of disease-causing rare variants. Lastly, our thorough phenotyping strategy allowed us to compare a relaxed vs. stringent case/control definition using ICD9/10, RxNorm, and CPT codes. As with many studies that rely on EHR data, the current study encountered limitations with regard to available phenotypic data. For example, no ophthalmology clinic notes, intraocular pressure measurements or ocular imaging data are currently available for *AoU* participants.

In summary, our multi-ancestry GWASs and meta-analyses of WGS in the ancestrally diverse *AoU* Research Program replicated known POAG loci and identified a novel locus in AFR individuals that replicates in an independent dataset and contains genes with disease-relevant biological functions. While additional fine-mapping and functional studies will be required to identify the causal variant(s) and gene(s) at this locus, the replication of rs76935404[T] in an independent clinically phenotyped African ancestry cohort supports the robustness of this association. Ongoing efforts to increase the size and diversity of multiple datasets with WGS will further improve our understanding of the genetic architecture of this common, blinding disease. Ultimately, this can lead to improved approaches to disease diagnosis, treatment, and prevention that will benefit all individuals.

## Data Availability

All of Us data are available on the cloud-based Researcher Workbench. The codes used for the analyses described in this manuscript are available upon request.

## Acknowledgements

We gratefully acknowledge *All of Us* Research Program participants for their contributions, without whom this research would not have been possible. We also thank the National Institutes of Health’s *All of Us* Research Program for making available the participant data and whole genome sequences examined in this study. The study presented in this manuscript was funded by the National Institutes of Health (K23EY035734 to IFA, 5T32EY007157 and 5T32HL007567 to LAC, R01EY022305 to JLW, R01EY033829 to JNCB, R01EY031424 to AVS, R01EY032559 to LRP and JLW, R01EY036460 to LRP, and P30EY014104 to Massachusetts Eye and Ear), Research to Prevent Blindness (Tom Wertheimer Career Development Award in Data Science to IFA), American Glaucoma Society (MAPS Award and Young Clinician-Scientist Grant to IFA), Prevent Blindness Ohio (Young Investigator Student Fellowship Award for Female Scholar in Vision Research to LAC), The Glaucoma Foundation (LRP), UK Research and Innovation Future Leaders Fellowship (MR/Y033930/1 to APK), Alcon Research Institute (Young Investigator Award to APK), Lister Institute for Preventive Medicine Award (APK), NIHR Biomedical Research Centre at Moorfields Eye Hospital (APK), and UCL Institute of Ophthalmology (APK).

---

**Supplementary Figure 1.**
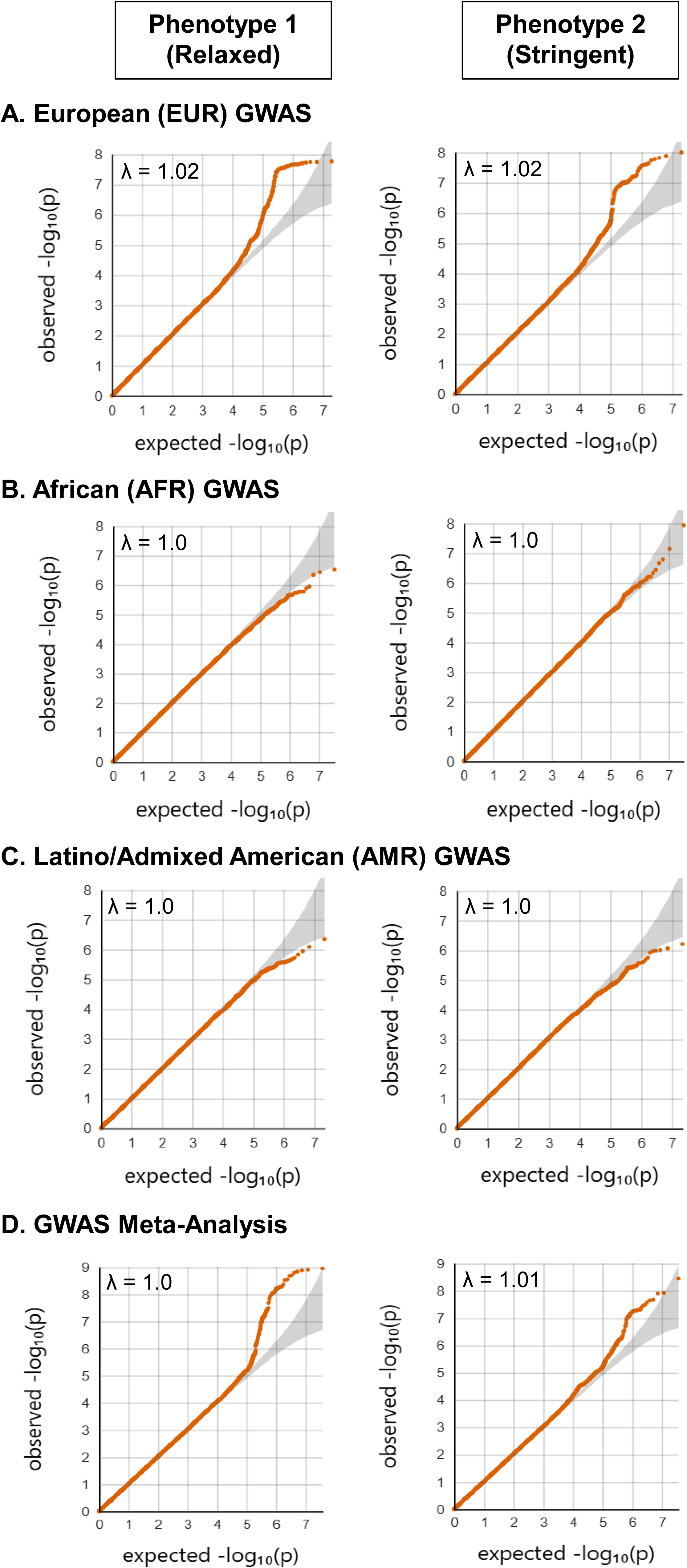
Quantile-quantile (QQ) plots for genome-wide association study (GWAS) results. The x-axis plots the expected -log_10_ p-value under the null hypothesis of no association, while the y-axis plots the observed -log_10_ p-value. The λ indicates the genomic inflation factor. There was no evidence of genomic inflation in the ancestry-stratified GWASs (A-C) or in the cross-ancestry meta-analyses (D) for either phenotype definition.

**Supplementary Figure 2.**
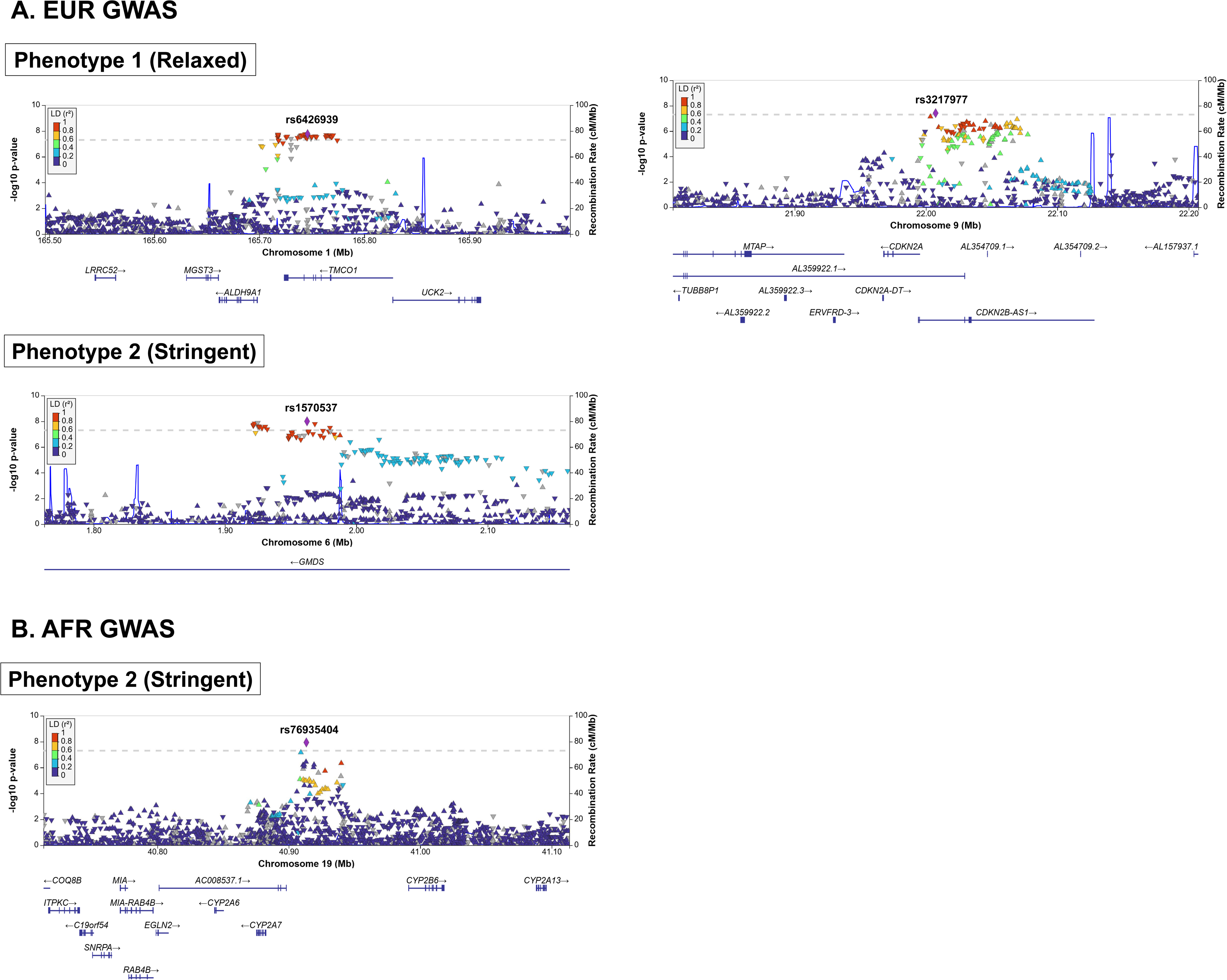
LocusZoom plots for genome-wide significant loci in the *All of Us* Research Program European (EUR) and African (AFR) genome-wide association studies (GWASs). All plots are mapped to genome build GRCh38. The reference variant for each plot (denoted by a purple diamond) is the lead POAG-associated variant in the genomic locus with available data in the 1000 Genomes Project. The linkage disequilibrium (LD) structures were generated using the 1000 Genomes Project reference panel corresponding to the population studied in each GWAS (EUR for EUR GWAS and AFR for AFR GWAS). No genome-wide significant loci were identified in the AFR GWAS using Phenotype 1 (relaxed) or in the AMR GWAS using either phenotype definition.

**Supplementary Figure 3.**
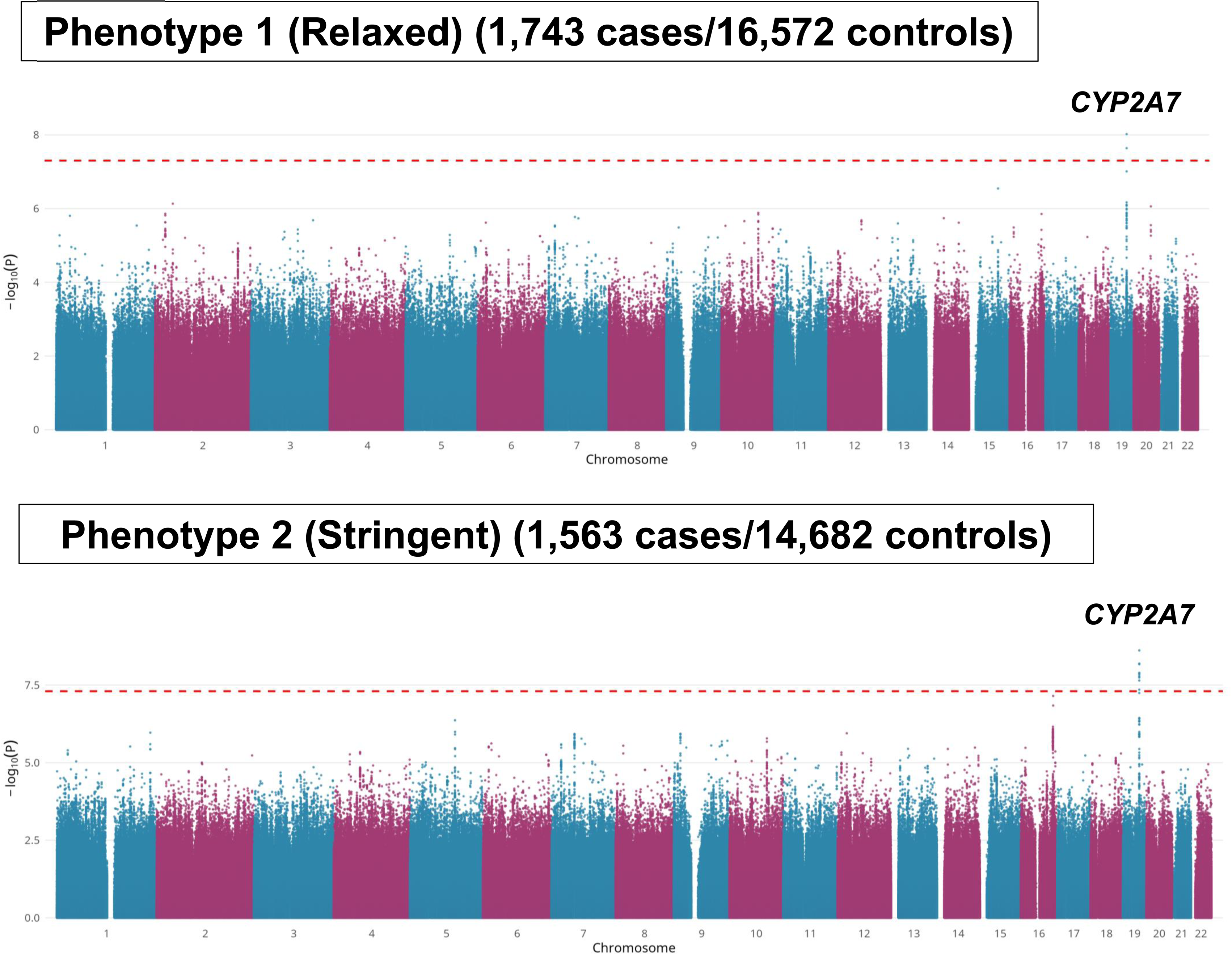
Manhattan plots for a meta-analysis of the *All of Us* Research Program African (AFR) genome-wide association studies (GWASs) with a GWAS from the subset of the Genetics of Glaucoma in People of African Descent (GGLAD) consortium enrolled at Duke University (Durham, NC, USA) (GGLAD-Duke). Primary open-angle glaucoma (POAG) GWASs were performed for the *All of Us* Research Program AFR cohorts using both the stringent and relaxed phenotype (Figure 2). The results were then meta-analyzed with results from an AFR POAG GWAS using the Duke University subset of participants from GGLAD (Hauser M.A. et al., *JAMA* 2019). The x-axis displays the chromosome where each variant is located, and the y-axis displays the -log_10_ p-value for association of the variant with POAG in the respective meta-analysis. The red dashed line represents the genome-wide significance threshold (p=5×10^-8^, -log_10_p = 7.30). The *CYP2A7* locus reached genome-wide significance in the meta-analysis with *All of Us* Phenotype 1 and retained significance when meta-analyzed with the Phenotype 2 discovery cohort.

**Supplementary Figure 4.**
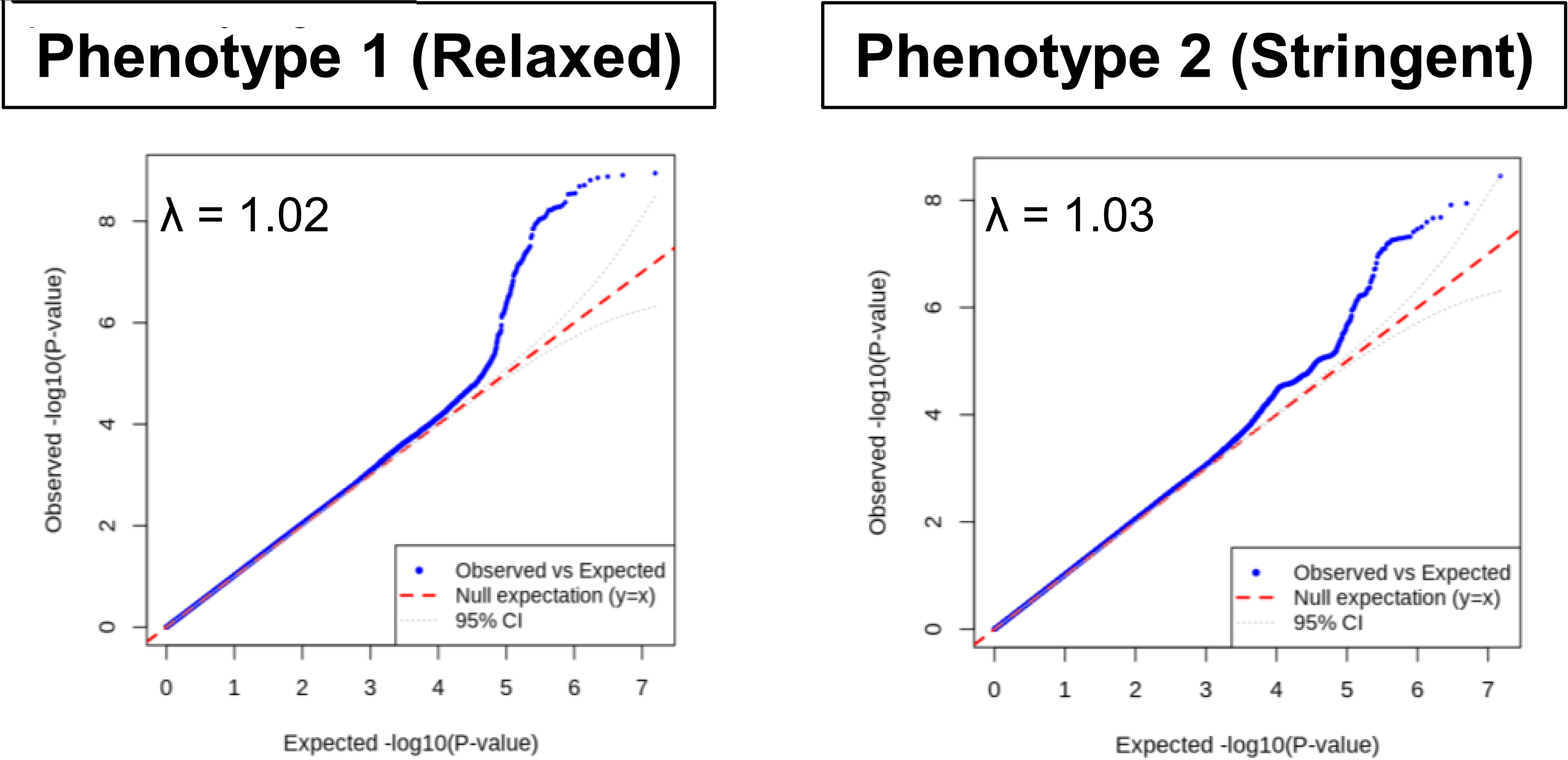
Quantile-quantile (QQ) plots for meta-analyses of the *All of Us* Research Program African (AFR) genome-wide association studies (GWASs) with a GWAS from the subset of the Genetics of Glaucoma in People of African Descent (GGLAD) consortium enrolled at Duke University (Durham, NC, USA) (GGLAD-Duke). The x-axis plots the expected -log10 p-value under the null hypothesis of no association, while the y-axis plots the observed -log10 p-value. The λ indicates the genomic inflation factor. There was no evidence of genomic inflation in the meta-analyses for either phenotype definition.

**Supplementary Figure 5.**
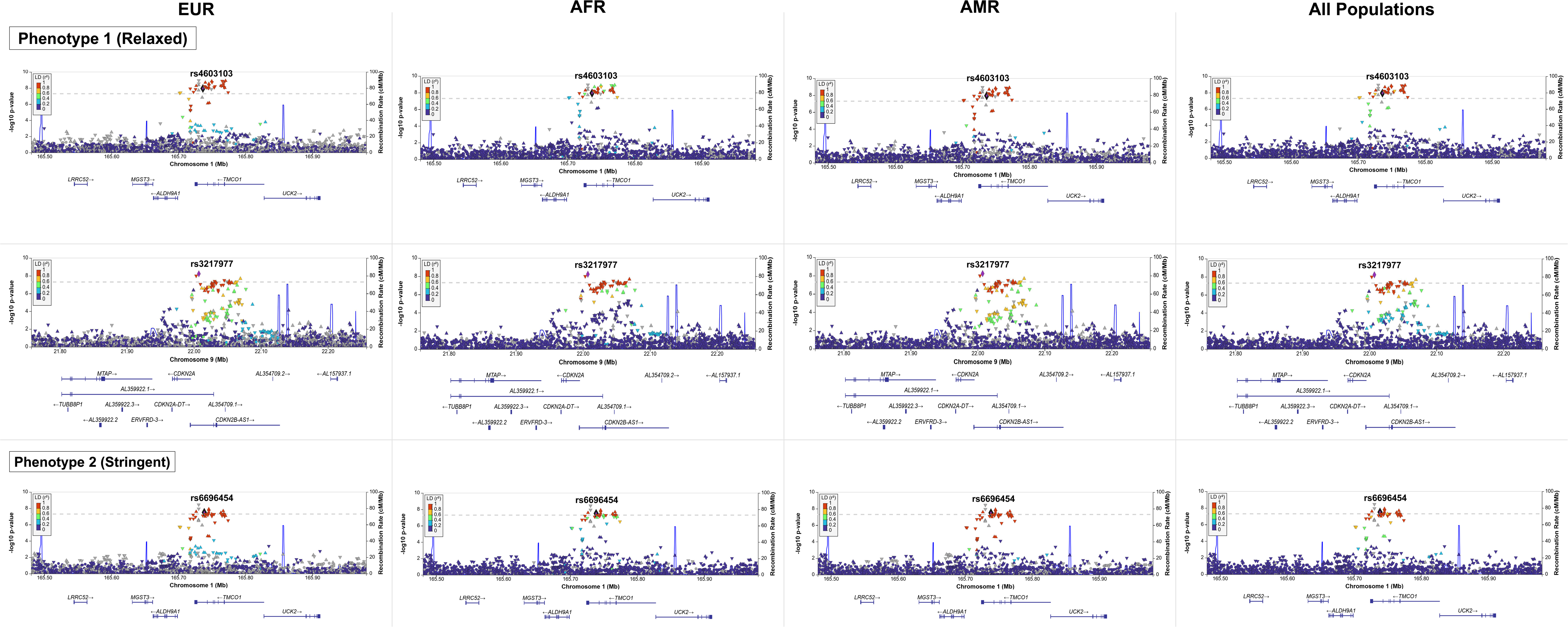
LocusZoom plots for genome-wide significant loci in the *All of Us* Research Program cross-ancestry meta-analyses. All plots are mapped to genome build GRCh38. The lead POAG-associated variant with available data in the 1000 Genomes Project in each genomic locus is labeled. For each of the genome-wide significant loci, linkage disequilibrium (LD) structures are displayed for European (EUR), African (AFR), Latino/Admixed American (AMR), and all five super-populations in the 1000 Genomes Project (EUR, AFR, AMR, East Asian, and South Asian) combined.

**Supplementary Figure 6.**
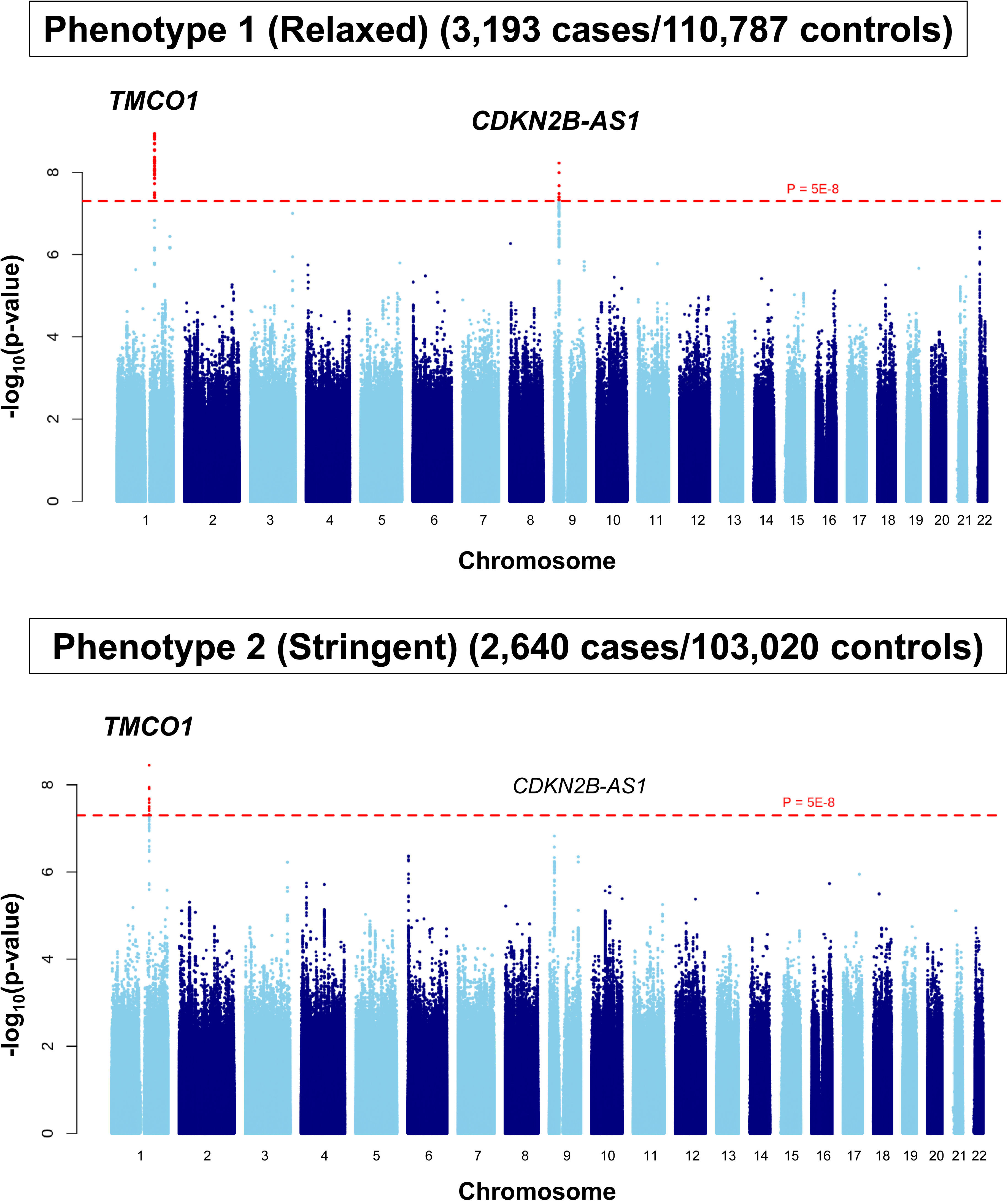
Manhattan plots for the *All of Us* Research Program cross-ancestry primary open-angle glaucoma (POAG) genome-wide (GWAS) meta-analyses using random effects models for each phenotype definition (relaxed and stringent). GWASs were performed for the European (EUR), African (AFR), and Latino/Admixed American (AMR) cohorts using a relaxed and stringent phenotype (Figure 2) and the results were meta-analyzed using a random effects model. The x-axis displays the chromosome where each variant is located, and the y-axis displays the -log_10_ p-value for association of the variant with POAG in the respective cross-ancestry meta-analysis. The red dashed line represents the genome-wide significance threshold (p=5×10^-8^, -log_10_p = 7.30). For variants reaching genome-wide significance, the nearest gene to the lead variant in the locus is labeled and bolded. The nearest gene(s) to genome-wide significant loci in one phenotype definition are also labeled on the Manhattan plot for the second phenotype definition. The results for this analysis are consistent with the fixed effects model (Figure 3).

**Supplementary Figure 7.**
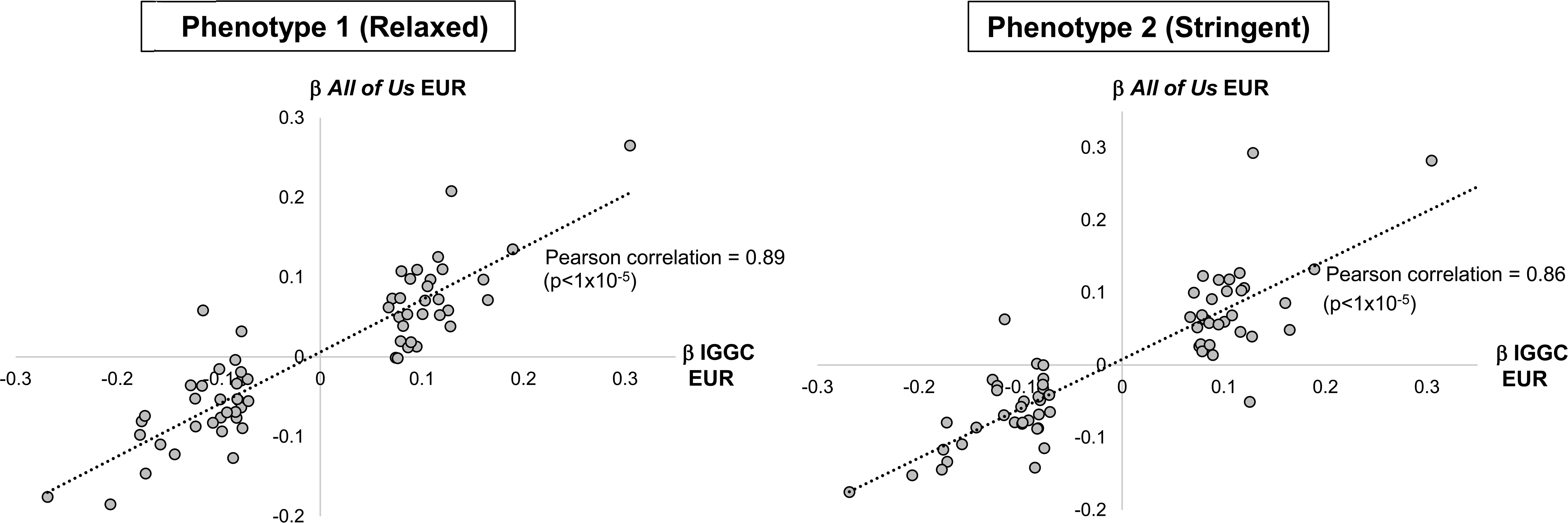
Correlation of effect sizes for known primary open-angle glaucoma (POAG)-associated variants identified in European (EUR) populations with their effect sizes in the *All of Us* Research Program EUR genome-wide association studies (GWASs) using each phenotype definition (stringent and relaxed). Correlation analyses were performed for all POAG-associated variants identified in the International Glaucoma Genetics Consortium (IGGC) EUR GWAS meta-analysis (16,677 cases/199,580 controls; 68 variants) (Gharahkhani P. et al., *Nature Communications* 2021). Each dot represents a specific variant. The beta regression coefficient (ß) from the IGGC EUR meta-analysis are plotted on the x-axis, while the ß from the *All of Us* Research Program EUR GWAS are plotted on the y-axis.

**Supplementary Figure 8.**
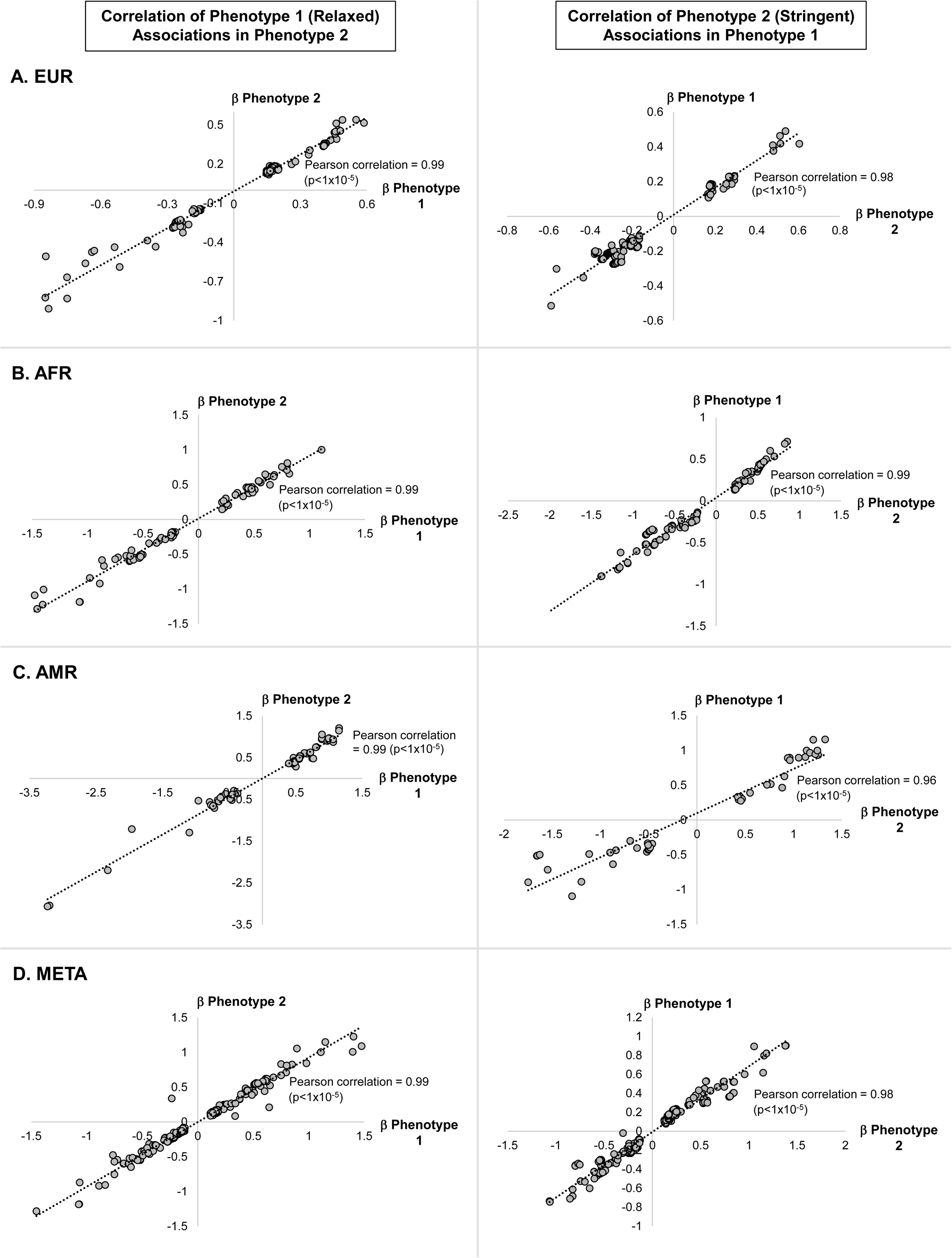
Correlation of effect sizes for top (p<1×10^-5^) loci identified in a genome-wide association study (GWAS) using one phenotype definition with effect sizes in the GWAS using the second phenotype definition. Correlation analyses using beta regression coefficients (ß) were performed for top variants for the *All of Us* Research Program European (EUR), African (AFR), Latino/Admixed Race (AMR), and cross-ancestry meta-analysis using each phenotype definition (relaxed and stringent). Each dot represents a specific variant.

**Supplementary Table 1.**
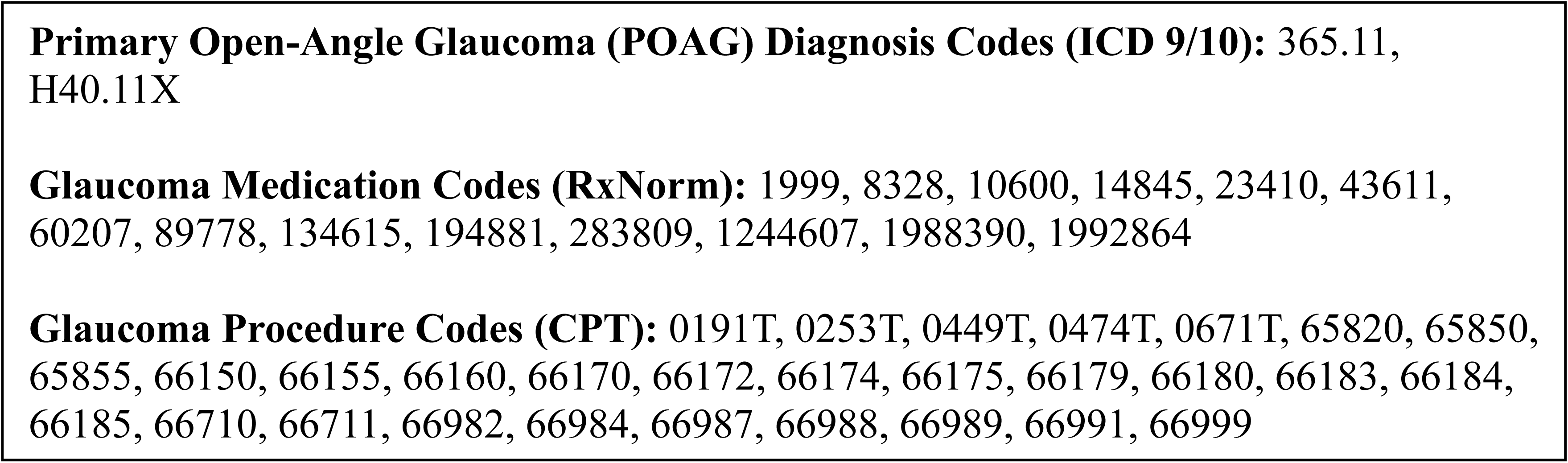
ICD 9/10, RxNorm, and CPT Codes Used to Generate Cohorts.

|  |
| --- |
| **Primary Open-Angle Glaucoma (POAG) Diagnosis Codes (ICD 9/10):** 365.11, H40.11X |
| **Glaucoma Medication Codes (RxNorm):** 1999, 8328, 10600, 14845, 23410, 43611, 60207, 89778, 134615, 194881, 283809, 1244607, 1988390, 1992864 |
| **Glaucoma Procedure Codes (CPT):** 0191T, 0253T, 0449T, 0474T, 0671T, 65820, 65850, 65855, 66150, 66155, 66160, 66170, 66172, 66174, 66175, 66179, 66180, 66183, 66184, 66185, 66710, 66711, 66982, 66984, 66987, 66988, 66989, 66991, 66999 |

**Supplementary Table 2.** Demographic characteristics of participants included in the All of Us genome-wide association analyses (by phenotype definition).

|  | Ancestry | Cases<br>(n) | Female n<br>(% of total) | Age<br>(mean ± SD) | Controls<br>(n) | Female n<br>(% of total) | Age (mean ±<br>SD) | *P*-value<br>(sex) | *P*-value<br>(age) |
| --- | --- | --- | --- | --- | --- | --- | --- | --- | --- |
| Phenotype 1<br>(Relaxed) | AFR | 1042 | 604 (58.0) | 69.4 ± 9.5 | 15966 | 8941 (56.0) | 71.0 ± 5.5 | 0.228 | 9.21×10<sup>-8</sup> |
|  | AMR | 305 | 174 (57.0) | 69.9 ± 10.5 | 10167 | 6100 (60.0) | 71.8 ± 5.8 | 0.329 | 1.82×10<sup>-3</sup> |
|  | EUR | 1846 | 886 (48.0) | 75.8 ± 9.1 | 84654 | 46560 (55.0) | 74.3 ± 6.5 | 2.54×10<sup>-9</sup> | 2.62×10<sup>-12</sup> |
|  | META | 3193 | 1660 (52.0) | 73.1 ± 9.9 | 110787 | 62041 (56.0) | 73.6 ± 6.4 | 7.37×10<sup>-6</sup> | 4.58×10<sup>-3</sup> |
| Phenotype 2<br>(Stringent) | AFR | 862 | 475 (55.1) | 71.2 ± 9.3 | 14076 | 7832 (55.6) | 72.7 ± 5.3 | 0.785 | 3.17×10<sup>-6</sup> |
|  | AMR | 250 | 127 (50.8) | 72.1 ± 10.2 | 9668 | 5802 (60.0) | 73.4 ± 5.6 | 3.36×10<sup>-3</sup> | 4.58×10<sup>-2</sup> |
|  | EUR | 1528 | 682 (44.6) | 77.2 ± 9.1 | 79276 | 42729 (53.9) | 75.6 ± 6.2 | 1.25×10<sup>-6</sup> | 1.12×10<sup>-11</sup> |
|  | META | 2640 | 1284 (44.2) | 74.7 ± 9.7 | 103020 | 56363 (53.5) | 75.0 ± 6.2 | 3.79×10<sup>-5</sup> | 0.114 |

*P*-values were calculated using the $\chi^2$ statistic for sex distribution and the Welch's t-test for age distribution. SD: standard deviation; AFR: African ancestry; AMR: Admixed American/Latino ancestry; EUR: European ancestry; META: cross-ancestry meta-analysis

**Supplementary Table 3.** Association of the novel genome-wide significant variant from the *All of Us* AFR Phenotype 2 (Stringent) POAG GWAS (rs76935404[T] near *CYP2A7*) across all ancestry-specific and cross-ancestry meta-analyses.

| GWAS | Cases/Controls | Effect Allele Frequency | OR | 95% CI | p-value |
| --- | --- | --- | --- | --- | --- |
| AFR | 1,042/15,966 | 0.29 | 1.28 | 1.16-1.41 | 4.5E-07 |
| EUR | 1,846/84,654 | 0.36 | 1.00 | 0.94-1.07 | 0.95 |
| AMR | 305/10,167 | 0.42 | 1.05 | 0.90-1.24 | 0.53 |
| META (fixed effects) | 3,193/110,787 | 0.35 | 1.09 | 1.03-1.15 | 2.0E-03 |
| META (random effects) | 3,193/110,787 | 0.35 | 1.09 | 1.04-1.13 | 2.0E-03 |

Phenotype 2 (Stringent)
| GWAS | Cases/Controls | Effect Allele Frequency | OR | 95% CI | p-value |
| --- | --- | --- | --- | --- | --- |
| AFR | 862/14,076 | 0.29 | 1.35 | 1.22-1.50 | 1.2E-08 |
| EUR | 1,528/79,276 | 0.35 | 1.00 | 0.93-1.08 | 0.95 |
| AMR | 250/9,668 | 0.41 | 1.12 | 0.94-1.34 | 0.22 |
| META (fixed effects) | 2,640/103,020 | 0.35 | 1.11 | 1.05-1.18 | 3.1E-04 |
| META (random effects) | 2,640/103,020 | 0.35 | 1.11 | 0.92-1.35 | 3.1E-04 |

POAG: primary open-angle glaucoma; GWAS: genome-wide association study; AFR: African; EUR: European; AMR: Latino/Admixed American; OR: odds ratio; CI: confidence interval

